# Comprehensive Druggable Genome-Wide Mendelian Randomization Reveals Therapeutic Targets for Kidney Diseases

**DOI:** 10.1101/2024.06.01.24308209

**Authors:** Zhihang Su, Qijun Wan

## Abstract

**Background:** Kidney diseases, including membranous nephropathy (MN), IgA nephropathy (IgAN), and chronic kidney disease (CKD), pose significant global health challenges due to their high prevalence and severe outcomes. There is still an urgent need to discover new targets for treating kidney diseases. Mendelian randomization (MR) has been widely used to repurpose licensed drugs and discover novel therapeutic targets. Thus, we aimed to identify novel therapeutic targets for Kidney diseases and analyze their pathophysiological mechanisms and potential side effects.

**Methods:** Integrated with currently available druggable genes, Summary-data-based MR (SMR) analysis was conducted to estimate the causal effects of blood expression quantitative trait loci (eQTLs) on kidney diseases. A study was replicated using distinct blood eQTL and diseases genome-wide association study (GWAS) data sources to validate the identified genes. The eQTL data was obtained from eQTLGen and GTEx v8.0, with sample sizes of 31,684 and 15,201, respectively. The data on kidney diseases was sourced from the Kiryluk Lab, CKDgen, and the Finngen consortium, with sample sizes ranging from 7,979 to 412,181. Subsequently, reverse two-sample MR and colocalization analysis were employed for further validation. Finally, the potential side effects of the identified key genes in treating kidney diseases were assessed using phenome-wide MR and mediation MR.

**Results:** After correcting for the false discovery rate, a total of 20, 23, and 6 unique potential genes were found to have causal relationships with MN, IgAN, and CKD, respectively. Among them, MN showed validated associations with one gene (HCG18), IgAN demonstrated associations with four genes (AFF3, CYP21A2, DPH3, HLA-DRB5), and chronic kidney disease (CKD) displayed an association with one gene (HLA-DQB1-AS1). Several of these key genes are druggable genes. Further phenome-wide MR analysis revealed that certain genes may be associated with diabetes, fat metabolism, and infectious diseases, suggesting that these factors could potentially serve as mediators.

**Conclusions:** This study presents genetic evidence that supports the potential therapeutic benefits of targeting these key genes for treating kidney diseases. This is significant in prioritizing the development of drugs for kidney diseases.

## 1 Introduction

Kidney diseases, including membranous nephropathy (MN), IgA nephropathy (IgAN), and chronic kidney disease (CKD), pose significant global health challenges due to their high prevalence and severe outcomes. MN, characterized by the deposition of immune complexes in the glomerular basement membrane, often leads to nephrotic syndrome and progressive renal failure if untreated(1). IgAN, the most common glomerulonephritis worldwide, is marked by the accumulation of IgA in the glomeruli, causing inflammation and potentially advancing to end-stage renal disease (ESRD)(2). CKD, defined by the gradual loss of kidney function over time, affects millions and significantly increases the risk of cardiovascular disease and mortality(3). Despite advancements in understanding the pathophysiology of these diseases, effective treatments remain limited. Current therapeutic strategies primarily focus on managing symptoms and slowing disease progression, but they do not halt or reverse kidney damage. For instance, the treatment of MN and IgAN often involves immunosuppressive therapies, which can have substantial side effects and variable efficacy. Similarly, CKD management revolves around controlling hypertension and diabetes, delaying progression rather than curing the disease. The identification of novel therapeutic targets is crucial to developing more effective treatments. With advances in genomics, proteomics, and bioinformatics, there is an unprecedented opportunity to uncover molecular pathways involved in kidney disease pathogenesis. Targeting these pathways could lead to innovative therapies that address the underlying causes of kidney damage rather than merely alleviating symptoms. This approach holds promise for improving patient outcomes and quality of life, highlighting the importance of ongoing research in this field.

Mendelian randomization (MR) is a powerful epidemiological method that leverages genetic variants as instrumental variables to assess the causal effects of modifiable risk factors on disease outcomes(4). By utilizing the random assortment of genes from parents to offspring, MR mimics the conditions of a randomized controlled trial, thereby minimizing confounding and reverse causation issues that often plague observational studies(5,6). This technique has gained prominence in identifying potential drug targets by providing insights into the causal relationships between biomarkers, risk factors, and disease outcomes(7,8).

In recent years, MR has been increasingly applied to drug discovery and development. By correlating genetic variants that influence drug target genes with clinical outcomes, MR can validate the therapeutic potential of specific targets before costly clinical trials are initiated. For instance, the use of MR has facilitated the identification of novel therapeutic targets for cardiovascular diseases by confirming the causal role of lipid levels and inflammatory markers in disease progression(9,10). Similarly, MR studies have highlighted the potential of targeting specific metabolic pathways in treating diabetes and other metabolic disorders(11,12).

The application of MR in drug target discovery is not without challenges, including the need for robust genetic instruments and comprehensive datasets. However, integrating MR with advanced genomic technologies and large-scale biobanks holds great promise for the future of precision medicine. By enhancing our ability to identify and validate causal relationships between potential drug targets and disease outcomes, MR can accelerate the development of effective and safe therapeutics, ultimately improving patient care.

## 2 Methods

This study was conducted according to the reporting guidelines outlined by the Strengthening the Reporting of Observational Studies in Epidemiology (STROBE) initiative.

### 2.1 Identification of druggable genes

The information regarding druggable genes was obtained from the Drug-Gene Interaction Database (DGIdb V.5.0.6) and the latest review on the “druggability” of genes(13). DGIdb provides comprehensive information on drug-gene interactions and druggable genes from various sources, including publications, databases, and other web resources(14). For this study, we retrieved the “categories” data from DGIdb, which was released in February 2022. This dataset contains a comprehensive list of druggable genes mapped to Entrez genes in DGIdb. Furthermore, we supplemented our findings with additional gene listings of druggable genes from the review conducted by Finan et al.

### 2.2 Expression quantitative trait loci

The blood eQTL dataset used in this study was obtained from eQTLGen (https://eqtlgen.org/). The dataset comprises blood samples from 31,684 individuals of European ancestry who were in good health(15). It covers cis-eQTLs of 16,987 genes. We exclusively considered cis-eQTL results that exhibited high significance (false discovery rate (FDR) < 0.05) and also incorporated allele frequency information. We identified genetic variants located within a 1000 kb range on both sides of the coding sequences (cis) that exhibited a strong association with gene expression. These variants were extracted using eQTL summary statistics obtained from the eQTLGen Consortium. Nonetheless, it should be noted that eQTLGen does not encompass variants linked to gene expression levels on the X and Y chromosomes, as well as mtDNA. Further details regarding the data can be found in the original publication. Additionally, to validate the obtained results, we employed the whole-blood cis-eQTL data from the Genotype-Tissue Expression (GTEx) project, specifically GTEx V.8 (https://gtexportal.org/home/datasets), which consisted of 15,201 samples.

### 2.3 Kidney diseases data

The primary source of discovery data for this study is the MN and IgAN databases provided by the Kiryluk Lab(16,17), along with the CKD database provided by the Finngen Consortium. To validate our findings, we additionally utilized the databases from the Finngen Consortium, CKDgen, and other sources. Regarding the Kiryluk Lab data, all MN cases were diagnosed utilizing the gold standard method of renal biopsy, whereas all IgAN cases were defined through dominant mesangial IgA staining on renal biopsy immunofluorescence. Cases potentially associated with secondary causes (e.g., Hepatitis, autoimmune, or malignant diseases) were excluded to uphold study rigor.

CKDgen’s research covers chronic kidney disease (CKD), which is defined by the criteria of eGFRcrea < 60 ml/min/1.73m^2^ and includes conditions such as urinary albumin-to-creatinine ratio (UACR > 30 mg/g) and gout(18,19). As the study samples are predominantly sourced from the general population, the research did not primarily focus on severe CKD and severe proteinuria. The analysis concentrates on cases with eGFR > 15 ml/min/1.73m^2^ and excludes patients receiving dialysis treatment.

### 2.4 Mendelian randomization

The MR method necessitates the fulfillment of three fundamental assumptions. To broaden the scope of MR, we performed a summary-data-based MR approach that estimates pleiotropic associations between genetic determinants of traits (e.g., gene expression, DNA methylation, or protein abundance as exposures) and our target complex traits, including MN, IgAN, and CKD. The heterogeneity-independent instruments (HEIDI) test is one of the sensitivity analysis techniques utilizing an external reference to estimate linkage disequilibrium (LD). Following false discovery rate (FDR) correction, genes exhibiting SMR < 0.05 and HEIDI test > 0.05 are deemed to possess causal relationships. Furthermore, we examine the observed effect sizes (OR values) to discern their roles as risk or protective factors. Subsequent to the completion of the SMR analysis, we conducted additional sensitivity analysis employing the Two-sample MR (TSMR) approach, encompassing inverse variance weighted (IVW) and the Wald ratio. Each method relies on distinct assumptions regarding the validity of instrumental variables to compute estimates of causal effects, thus furnishing robust evidence for our findings. In conducting TSMR, we employ data from the 1000 Genomes project (HG19/GRCh37) as a substitute in cases where effect allele frequency data is missing.

### 2.5 Phenome-wide MR and mediation MR

Using data from the UK Biobank as the outcome and key genes as the exposure, we employed phenome-wide Mendelian randomization (MR) to identify phenotypes associated with the key genes, potentially indicating comorbidity relationships with kidney diseases. Moreover, to identify intermediary factors associated with kidney diseases, we conducted two-sample MR analysis using key genes as the exposure, UK Biobank data as the mediator, and kidney diseases as the outcome.

### 2.6 Bayesian colocalization analysis

Occasionally, a single nucleotide polymorphism (SNP) may be situated within multiple gene regions. In such instances, if the SNP contains information on the expression of quantitative trait loci (QTL) for two or more distinct genes, its influence on kidney diseases (MN, IgAN, and CKD) will reflect a combination of various genes. Co-localization analysis is employed to validate the presence of shared causal genetic variations between kidney diseases and eQTL. Briefly, we conducted the colocalization analysis to investigate the co-localization of kidney disease risk and SNP within ±100 kb of the transcription start site (TSS) of each gene in eQTL, focusing on significant MR (Mendelian randomization) results from the discovery phase (P1 = 1×10^−5^, P2 = 1×10^−5^, and P12 = 1×10^−5^). P1 denotes the probability of a significant eQTL association with the provided SNP, P2 signifies the probability of the given SNP being associated with kidney disease, and P12 indicates the probability of the given SNP being associated with both kidney disease and eQTL outcomes. The researchers evaluated five hypotheses, and posterior probabilities (PP) are utilized to quantify the level of support for each hypothesis, categorized as PPH0 to PPH4, with a specific emphasis on PPH3 and PPH4. PPH3 represents the co-localization of gene expression and kidney disease, demonstrating distinct causal variations within the same gene locus, whereas PPH4 indicates the co-localization of gene expression and kidney disease, manifesting shared causal variations. Due to the limited power of co-localization analysis, we confined our analysis to genes that surpassed a threshold of PPH3+PPH4 ≥ 0.5 (0.5 to 0.8 indicating moderate strength, >0.8 indicating high strength).

### 2.7 Functional Enrichment Analysis

The functions of genes and potential signaling pathways associated with tumorigenesis and progression in kidney diseases were explored using the Metascape database (v3.5.20240101). By utilizing these selected functional sets in Metascape, our aim was to gain insights into the molecular functions, biological processes, pathways, and structural complexes associated with the genes in our dataset. Additionally, the Signature Module sets enabled us to explore cellular signatures associated with cell types, chemical and genetic perturbations, immunological signatures, and oncogenic signatures. The Miscellaneous sets provided information on transcription factor targets and gene-disease associations sourced from trusted databases, including TRRUST, PaGenBase, and DisGeNET. Furthermore, the L1000 sets allowed us to examine the effects of genetic perturbations, compounds, cDNA, and ligands. Lastly, we explored the vaccine response signatures, with a particular focus on COVID-related signatures, to gain insights into the immune response associated with our target genes. Through this comprehensive analysis, our aim was to uncover potential functional mechanisms, pathways, and interactions relevant to our target of interest. Gene Ontology (GO) enrichment analysis and Kyoto Encyclopedia of Genes and Genomes (KEGG) analysis were conducted. All genes that exhibited significant alterations after correction were included in this analysis. The GO analysis was categorized into three sections: BP (biological process), CC (cellular component), and MF (molecular function). A P-value below 0.05 was considered statistically significant.

## 3 Results

We successfully identified multiple causal associations between gene expression and kidney diseases in the blood through SMR tests. To control for type I errors across the entire genome, we conducted false discovery rate (FDR) correction, which revealed robust evidence of associations (FDR < 0.05). Subsequently, we performed HEIDI tests (P > 0.05) to explore whether these associations resulted from shared causal variants rather than pleiotropy. To support our findings, we conducted sensitivity analyses using other MR methods that are based on similar assumptions, demonstrating consistent outcomes. Additionally, to address confounding factors, we conducted a colocalization analysis, which assessed the posterior probability of shared causal variation (PPH4) between gene expression and kidney diseases, employing a threshold of >0.50. Additionally, we replicated our findings by performing additional analyses utilizing other GWAS studies. The identified genes were classified into four distinct target groups based on SMR, TSMR, colocalization analysis, and external replication results.

Genes with an FDR-corrected SMR-P value < 0.05 and a HEIDI test result > 0.05 were considered essential criteria and classified as tier 1 targets when supported by TSMR, colocalization, and replication. Genes supported by any two of TSMR, colocalization, and replication were classified as tier 2 targets. Genes supported by any one of TSMR, colocalization, or replication were classified as tier 3 targets. The remaining genes were classified as tier 4 targets.

Following the aforementioned principles, we identified a primary target (HCG18) and multiple secondary targets associated with MN. Moreover, we identified four primary targets (AFF3, CYP21A2, DPH3, HLA-DRB5) and several secondary targets associated with IgAN. Additionally, we identified various secondary targets associated with CKD (Table 1). In the case of MN, a decrease of 1 standard deviation (SD) in HCG18 expression was significantly associated with an 82% reduction in risk (OR: 0.18, 95% CI: 0.08-0.43, P^SMR^ = 9.79×10^-5^). Regarding IgAN, an increase of 1 SD in AFF3 expression was linked to a 46% reduced risk (OR: 0.54, 95% CI: 0.39-0.74, P^SMR^ = 1.64×10^-4^). Similarly, an increase of 1 SD in CYP21A2 expression was associated with a 42% reduced risk (OR: 0.58, 95% CI: 0.45-0.76, P^SMR^ = 5.53×10^-5^), an increase of 1 SD in DPH3 expression was correlated with a 27% reduced risk (OR: 0.73, 95% CI: 0.66-0.80, P^SMR^ = 3.89×10^- 11^), and an increase of 1 SD in HLA-DRB5 expression was linked to a 26% reduced risk (OR: 0.74, 95% CI: 0.43-0.86, P^SMR^ = 8.54×10^-5^). The reverse TSMR analysis revealed an inverse relationship between IgAN and only HLA-DPA1, HVCN1, and EHMT2, while no associations were found for the remaining targets in kidney diseases.

### 3.1 Phewas

Through comprehensive phenotypic Mendelian randomization, we uncovered a range of diseases and phenotypes associated with kidney disease. These diseases and phenotypes may exhibit comorbidity with kidney disease. Moreover, employing a two-step approach, we identified diabetes, lipid metabolism, and other factors as potential pivotal mediators that influence kidney disease via genetic pathways.

### 3.2 Functional Enrichment Analysis

Pathways associated with key genes were identified through functional enrichment analysis. The AKT gene, CD4-positive dendritic cells, the immunoreceptor signaling pathway (UPG0:0002429), SATO-mediated methylation-induced silencing in pancreatic cancer, KRIGE’s response to TOSEDOSTAT after 24 hours, and the carboxylic acid metabolism process may be associated with MN through key genes. Th17 cell differentiation and lipid metabolism may be associated with IgAN through key genes. Differences or upregulation between TARTE cells, plasma cells, and plasmablasts under specific conditions or treatments may be associated with CKD.

## 4 Discussion

To the best of our knowledge, this study represents the pioneering attempt to integrate blood genomics data with SMR, TSMR, Bayesian colocalization, phenotype scanning, KEGG, GO pathway analysis, and PPI analysis methods. Multiple databases were utilized to uncover and validate the potential roles of specific genes in MN, IgAN, and CKD. Through genetic prediction, we established a causal relationship between HCG18 and MN, AFF3, CYP21A2, DPH3, HLA-DRB5 and IgAN, and HLA-DQB1-AS1 and CKD (the highest-level targets discovered for each disease are listed here). After conducting a series of analyses, we ultimately identified the aforementioned genes as high-priority potential drug targets for kidney diseases. According to the target principles mentioned earlier, we believe these genes can function as potential drug targets for the treatment of kidney diseases, and several of them are druggable.

By utilizing SMR and employing various validation methods, we have newly identified several promising genes, thus enhancing their potential as therapeutic intervention targets. Therefore, to determine innovative targets for drug interventions in kidney diseases, we conducted comprehensive analyses to assess the causal relationships of genes. Causal relationships identified through MR may involve reverse causality, horizontal pleiotropy, or genetic confounding induced by LD. Therefore, in this study, we employed IVs exhibiting strong correlations with the genes, utilized the HEIDI test to estimate LD, and excluded IVs that exhibited evidence of linkage disequilibrium during MR implementation. Reverse Mendelian randomization analysis uncovered bidirectional causal relationships between HLA-DPA1, HVCN1, EHMT2, and IgAN. Additionally, Bayesian colocalization was utilized to mitigate bias introduced by LD (24). Employing a posterior probability threshold of 0.5, it was deemed to possess moderate to high colocalization strength, and we identified all genes that exhibited SMR positivity. Nevertheless, these associations alone do not fully elucidate the connections between the identified genes and kidney diseases. Additionally, we performed phenotype scanning analyses of the instrumental variables utilized in the MR analysis to address confounding factors. Thus, the aforementioned genes have the potential to serve as drug targets for kidney diseases, with a particular focus on Tier 1 targets.

### Membranous Nephropathy

HLA Complex Group 18 (HCG18) is a long non-coding RNA gene. For HCG18 and MN, there are some indications of a potential relationship, although it is not well-established. Research has shown that long non-coding RNAs (lncRNAs), including HCG18, are involved in various biological processes and diseases, including cancer, COVID-19 infections, and potentially other complex conditions like nephropathy(20–22). However, specific studies directly linking HCG18 to MN are sparse. One study suggests that lncRNAs like HCG18 could regulate immune responses and cellular processes that might be relevant to diseases like MN, but conclusive evidence is lacking.

### IgA nephropathy

AFF3, also known as ALF Transcription Elongation Factor 3, is a protein-coding gene that encodes a nuclear transcriptional activation factor with tissue-restricted expression, primarily in lymphoid tissues. AFF3 is associated with diseases such as Familial Syndrome and Intellectual Developmental Disorder, specifically X-linked 109. Furthermore, it is suggested that this gene may contribute to lymphoid development and tumor formation. CYP21A2, also known as Cytochrome P450 Family 21 Subfamily A Member 2, is a protein-coding gene. CYP21A2 is associated with diseases such as Congenital Adrenal Hyperplasia due to 21-hydroxylase Deficiency and Classical Congenital Adrenal Hyperplasia, both caused by 21-hydroxylase deficiency. HLA-DRB5, a member of the Major Histocompatibility Complex Class II, is a protein-coding gene that encodes the DR Beta 5 subunit.

HLA-DRB5 is associated with diseases such as Pityriasis Rosea and Systemic Lupus Erythematosus. It involves related pathways, including TCR Signaling and Phosphorylation of CD3 and TCR zeta chains. DPH3, also called Diphthamide Biosynthesis 3, is a protein-coding gene. DPH3 is associated with diseases such as Diphtheria and Melanotic Schwannoma. It involves related pathways such as Protein Metabolism, γ-carboxylation, hypusine formation, and arylsulfatase activation.

### Chronic Kidney Disease

HLA-DQB1-AS1, an RNA gene classified as a long non-coding RNA (lncRNA), It has been suggested to be associated with Osteoarthritis and Muscle Atrophy.

Our study has several limitations. Firstly, the blood genomic data used in our analysis were sourced from two distinct studies that may have utilized varying measurement standards. However, we chose a database with a large sample size as the discovery cohort, and another one as the validation cohort. Secondly, during the validation process using different databases, certain genes did not produce results, resulting in missing data. Missing data does not imply insignificant findings; instead, it signifies unknown information. Thirdly, the data utilized in our experiments solely represent the European population, necessitating further research to extend the generalizability of our findings to other ethnicities. Additionally, the precise mechanisms underlying these genes and kidney diseases are still unknown. It is essential to acknowledge that unvalidated data analysis may encompass aspects necessitating experimental verification, thereby enabling the achievement of objectives beyond the scope of data analysis alone. Our data can be utilized in future research to validate drug targets. Enhancing our comprehension of the genetic regulation of drug targets and the circulating levels of biomarkers may improve drug interventions and clinical trials.

## Data Availability

All data were obtained from publicly available databases.

## 5 Data availability

eQTLs data was derived via public research(23), and MN data was retrieved from Kiryluk Lab(16). We used R language version 4.3.0 for our analysis. In R language, we utilized the “coloc (https://github.com/chr1swallace/coloc.git)” and “TwoSampleMR (https://github.com/MRCIEU/TwoSampleMR.git)” and “locuscompare” packages for our analysis(24).

## 6 Conflict of Interest

The authors declare that the research was conducted without any commercial or financial relationships that could potentially create a conflict of interest.

## 7 Ethic

The data utilized in this analysis were obtained from publicly accessible databases. All of the data have been de-identified and received ethical approval from the appropriate ethics committee.

Consequently, this study does not necessitate individual ethical approval.

## 8 Author Contributions

Zhihang Su and Qijun Wan jointly accomplished all efforts. Rui Xue, Jiaqi Liu, Liling Wu, and Yuan Cheng contributed to revising and refining the article.

## 9 Funding

Shenzhen Key Medical Discipline Construction Fund (SZXK009) and SZSM202211013.

## 10 Acknowledgments

Sincere thanks to all participants who participated in producing the public database.

## 12 Figure legends

**Figure 1:**
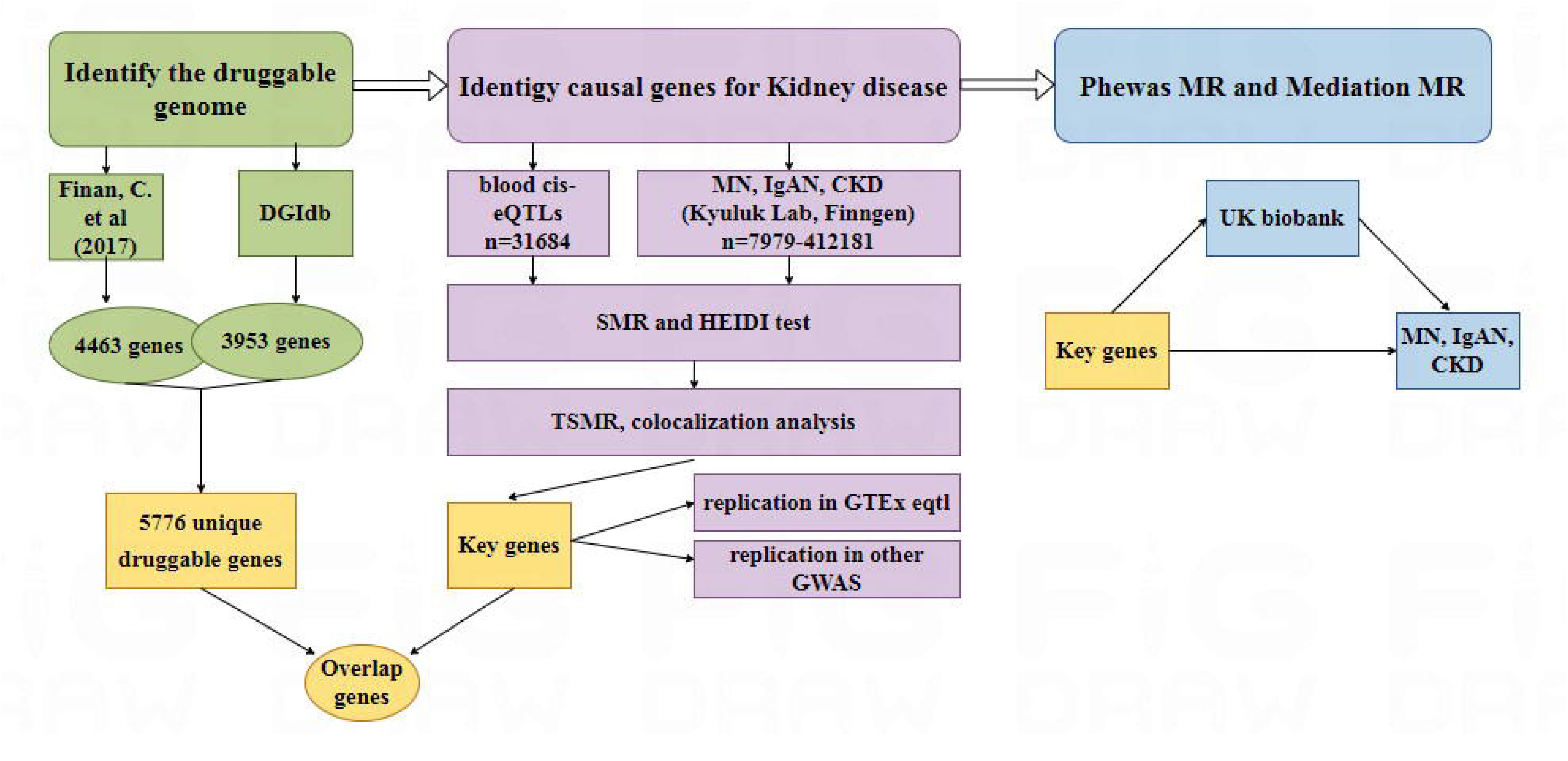
Study design. MN: membranous nephropathy; IgAN: IgA nephropathy; CKD: chronic kidney disease; HEIDI test: heterogeneity in dependent instruments; FDR: false discovery rate; PPH4: posterior probabilities of hypotheses 4.

## Notes

### Competing Interest Statement

The authors have declared no competing interest.

### Author Declarations

The data utilized in this analysis were obtained from publicly accessible databases. All of the data have been de-identified and received ethical approval from the appropriate ethics committee. Consequently, this study does not necessitate individual ethical approval.

